# Cage and experimental house trials to optimize the design of dried attractive bait stations for the control of *Aedes aegypti*

**DOI:** 10.1101/2025.11.05.25339578

**Authors:** Luis F. Quintanilla, Julieanne Miranda Bermudez, Grayson C. Brown, Nexilianne Borrero Segarra, Marco Neira, David A. Larsen

## Abstract

Dengue, Zika, and chikungunya, all transmitted by *Aedes aegypti* mosquitoes, present a significant and growing global health challenge. Contact-based insecticide such as pyrethroids, organophosphates, and organochlorines, are a core component of strategies to control *Ae. aegypti*. However, reliance on these chemical insecticides alone presents long-term challenges, particularly around increasing insecticide resistance. There are relatively few novel tools available that target adult mosquitoes through alternative mechanisms. Innovative, complementary approaches that can enhance the effectiveness and sustainability of existing mosquito control programs are urgently needed. Newly explored vector management tools are attractive toxic sugar baits (ATSBs), targeting the sugar-feeding behavior of both male and female mosquitoes, and allow for an ingested insecticide. Dried Attractive Bait Stations (DABS), a novel variant of ATSBs, have shown promising results for indoor *Ae. aegypti* control. We conducted cage and experimental house trials of DABS on laboratory-reared *Ae. aegypti* mosquitoes in Ponce, Puerto Rico. In cage trials, both male and female *Ae. Aegypti* that were exposed to DABS for 48 hours experienced near complete mortality, while control groups showed universal survival. All tested concentrations (1%, 2%, and 4% boric acid compounds) significantly reduced survival compared to controls (*p* < 0.001), with no significant differences between concentrations (*p* = 0.50). Water availability reduced DABS efficacy, lowering mortality by 47% in females (*p* < 0.001) and 32% in males (*p* < 0.001). In experimental house trials, DABS reduced mosquito survival by ∼75% (*p* < 0.001), with deployment of four DABS devices per room suggesting the greatest reduction in mosquito survival (*p* = 0.06–0.10). Optimal performance was observed at placing DABS 0.75 m above the floor (*p* < 0.001) and results suggest that medium-sized DABS may be more effective than larger (*p* = 0.09). Natural light increased mosquito survival (*p* = 0.028), but its interaction with DABS treatment was not significant (*p* = 0.322) and room-to-room variability was minimal. A notable limitation was the interference of ants during household trials, which led to the removal of mosquito carcasses and potential data loss, highlighting the need to further investigate alternative sugar sources and DABS interaction with non-target organisms. Despite these challenges, the results support DABS as a promising, scalable *Aedes aegypti* control tool. Future research should confirm effectiveness under varied field conditions, and assess the community acceptability for broader deployment.

## Introduction

The *Aedes aegypti* mosquito poses a significant global health threat as the primary vector of dengue, Zika, chikungunya, and yellow fever viruses. The global spread of *Ae. aegypti* has contributed to the rapid expansion of dengue, Zika, and chikungunya, with 56 million new dengue cases alone reported globally in 2019 (WHO, 2024) and over 13 million dengue cases in the Americas and Caribbean by 2024 (CDC, 2025). Dengue is now endemic in 128 countries (WHO, n.d.), including Puerto Rico in the United States, putting over 40% of the world’s population at risk (Hossain et al., 2021). Chikungunya and Zika viruses have also spread across the globe following their initial re-emergence (Grabenstein & Tomar, 2023; Noisumdaeng et al., 2023). The long-term and often indiscriminate use of contact-based chemical insecticides to control *Ae. aegypti* has led to the spread of resistance genes among *Ae. aegypti* populations (Rodríguez et al., 2007), reducing their effectiveness (Dusfour et al., 2019; Estep et al., 2017; Paeporn et al., 2005).

An emerging vector control strategy involves the development of attractive toxic sugar baits (ATSBs), which exploit the natural sugar-feeding behavior of adult *Ae. aegypti* by combining attractive sugar sources with toxic compounds designed to kill upon ingestion (Hossain et al., 2021). Any toxic compound that does not deter feeding on the ATSB is sufficient, and investigators have experimented with inorganic toxicants, biopesticides, pyrroles, and double-stranded RNA, among others (Fiorenzano et al., 2017). Inorganic toxicants stand out from the rest due to their very low cost, popular usage to control other insects like ants, low toxicity to vertebrates including humans, and limited evidence of resistance build up in mosquitoes (Murray, 1995; Pearson et al., 2020). Compared to the limited number of contact-based adulticides approved by the U.S. Environmental Protection Agency (EPA) for use within homes, there is an expanding list of inorganic toxicants with distinct modes of action that offer promising new directions for *Ae. aegypti* control (EPA, 2025). Ingestible inorganic toxicants exert their toxic effect by disrupting the integrity and function of the digestive tract, differing from the neurotoxic pathways targeted by contact-based insecticides (Cochran, 1995; Habes et al., 2006; Sippy et al., 2020).

Dried Attractive Bait Stations (DABS) are a novel variant of ATSBs and have shown promising results for indoor *Ae. aegypti* control. DABS utilize a dried sugar-toxic compound solution applied to a high-contrast (black-on-white) surface to attract both male and female mosquitoes. When *Ae. aegypti* land on the DABS, they detect the sugar using taste receptors located on their feet (Dennis et al., 2019) and ingest the toxic compound. Sippy et al. (2020) demonstrated the potential effectiveness of DABS in controlling *Ae. aegypti* through experimental house trials in Ecuador.

Given that *Ae. aegypti* are peridomestic mosquitoes that primarily live in and around human dwellings and enter homes in search of hosts for blood meals (Powell & Tabachnick, 2013), indoor deployment of DABS represents a strategic control approach. In addition to its entomological effectiveness, DABS offers practical advantages: it is affordable, simple to use, simple to make, easily distributed during a campaign, and discreetly placed without disturbing residents. Moreover, indoor placement of DABS would pose minimal to no risk to non-target organisms such as pollinators (Khallaayoune et al., 2013).

In this study, we performed cage and experimental house trials to optimize key parameters required for the implementation of a DABS-based strategy for controlling *Ae. aegypti* in real-world conditions, using boric acid as the toxic compound. Study parameters included device size, placement height, number of DABS devices per room, optimal concentration of boric acid, and the effect of DABS in male mosquitoes. The information gathered in these experiments will inform future field trials aimed at evaluating the efficiency of DABS as a tool for *Ae. aegypti* control.

## Methods

### Study setting

We conducted the study in Ponce, Puerto Rico. The municipality of Ponce, located in the southwestern region of Puerto Rico, experiences a predominantly hot and humid climate with and average rainfall of approximately 36.0 inches (920 mm) per year(CFWSC, 2016; CIMH, 2014). Ponce is the fourth-largest municipality on the island, with a population of approximately 137,000 residents (United States Census Bureau, 2020). The city faces significant socioeconomic challenges, with a median household income of around $20,000 and a poverty rate nearing 50% (DATAUSA, 2023; United States Census Bureau, 2020). In 2024, Puerto Rico experienced a sharp increase in dengue cases, prompting the declaration of an epidemic (Ware-Gilmore et al., 2025).

### Fabrication of dried attractive bait stations

DABS were constructed using a modified version of the process proposed by Sippy *et al*. (2020). Briefly, 2mm thick, nontoxic premium black foam sheets (Craft Unlimited®, Puerto Rico, USA) were cut into rectangular pieces 21.6 x 14 cm (301.3cm^2^) and soaked on each side for 15 minutes in a 10% (w/v) sucrose solution containing 1-4% boric acid (MilliporeSigma®, Puerto Rico, USA). Once soaked, the devices were left to dry overnight. For each experimental replicate, the DABS were manufactured the day prior. Control DABS were prepared simultaneously using the same procedure, but without the addition of boric acid to the sugar solution.

### Mosquito rearing

All *Ae. aegypti* used in our experiments belong to a strain originally collected in the city of Ponce, Puerto Rico, and maintained under insectary conditions at the CDC Dengue Branch with regular re-introduction using collected eggs from the field. Eggs (F_2_ generation) were hatched at the insectary of the Puerto Rico Vector Control Unit (PRVCU), under standard laboratory conditions: 12h/12h light/dark photoperiod, ∼26°C temperature, 80% relative humidity. Finely ground rabbit food pellets (SMALL WORLD®, St. Louis, MO, USA) were provided as food during the larval phase. Pupae were sexed based on morphological traits (USAID, 2019) and placed in single-sex rearing cages, where adults were allowed to emerge. A 10% sucrose solution was provided *ad libitum* to adults.

For semi-field trials, 4-5 days old adult mosquitoes were transferred to cardboard cups using a mouth aspirator and transported to the experimental house inside an insulated box. Mosquitoes were released into experimental rooms through a screened window. Temperature and humidity were recorded at both the insectary and trial house using portable weather devices, Kestrel® model 55000 FWL (Nielsen-Kellerman Co., Pennsylvania, USA).

### Cage Trials

Cage trials were conducted using mosquito rearing cages measuring 30W × 30D × 30H cm under controlled laboratory conditions. Each trial involved cages containing 40–50 adult *Ae. aegypti*. To promote bait ingestion, mosquitoes were starved for 24 hours prior to treatment by withholding access to the sucrose solution provided. A single DABS or control device was placed in each cage, either with or without a water source, depending on the specific trial conditions. Following treatment, mosquito mortality was recorded every 6 hours by counting the number of immobile mosquitoes on the cage floor.

#### DABS concentration trials

Optimal DABS concentration was evaluated using four rearing cages per trial and varying boric acid levels in the solution used to make DABS. The 1% DABS used 10g, 2% used 20g, 4% used 40g, and the control contained no boric acid. Each device was placed in a cage in combination with a 50ml flask of water covered at the opening with a folded paper napkin secured in place. The napkin acted as a wick, allowing mosquitoes to drink through capillary action. Observations were recorded every 6 hours for 48 hours. Four replicates were collected from this trial.

#### DABS water effect trials

To assess the effect of water availability during DABS treatment, separate trials were conducted for female and male *Ae. aegypti* mosquitoes using four rearing cages per trial. 1% concentration was selected for the female trials due to previous data from Sippy et al. (2020). The 2% concentration was selected for the male trials based on cage preliminary data indicating that this boric acid concentration might kill *Ae. aegypti* more rapidly than a 1% dose. To evaluate the impact of accessible water, one treatment cage and one control cage received a 50 ml flask of water fitted with a folded paper napkin as a water source. The remaining two cages (one treatment, one control) received no water source. Mosquito mortality was recorded every 6 hours over a 48-hour period, and each trial was replicated three times independently for males and females.

### Experimental house trials

Experimental house trials were conducted in a modified vacant house where four rooms were refurbished for the purposes of this study (Figure 1D). Each room was emptied and fully painted with a white zero VOC paint, ProMar 200 Zero VOC Interior Latex (Sherwin-Williams®, Cleveland, Ohio, USA). There were also window screens installed in each room, which had between 1-2 windows each. Each room had one door leading to the hallway. The door connected to the room was a screen door with attached screen windows for mosquito release. Reared mosquitoes were starved for 24 hours by removing the sucrose solution from their cages. They were then transported in cardboard cups and released into each room through the screen door windows. Each room contained a source of water (i.e. wet paper towel). Treatments followed a Latin-square design across four replicates. After 48 hours, dead and alive mosquitoes were collected using a Prokopack Aspirator™ (John W. Hock Company®, Gainesville, Florida, USA).

**Figure 1.**
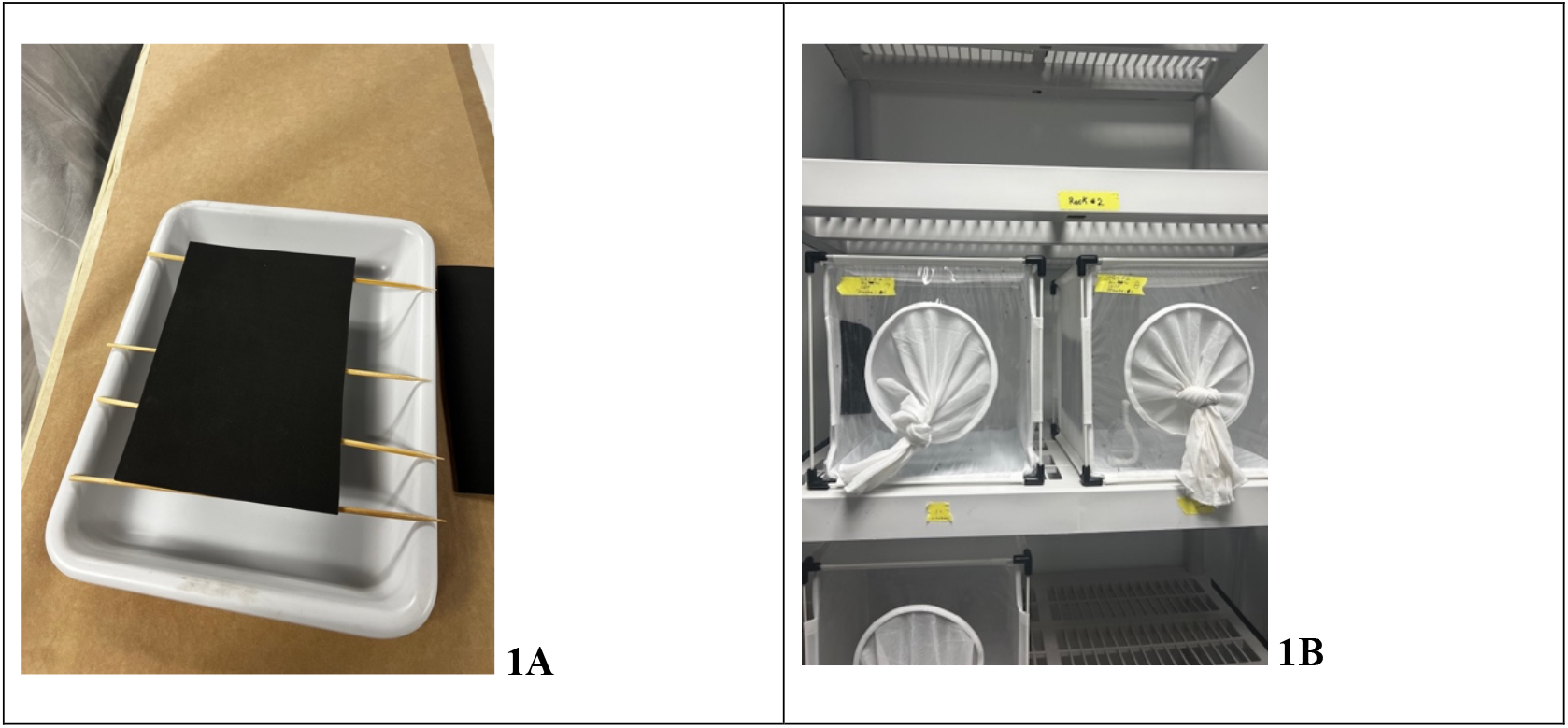
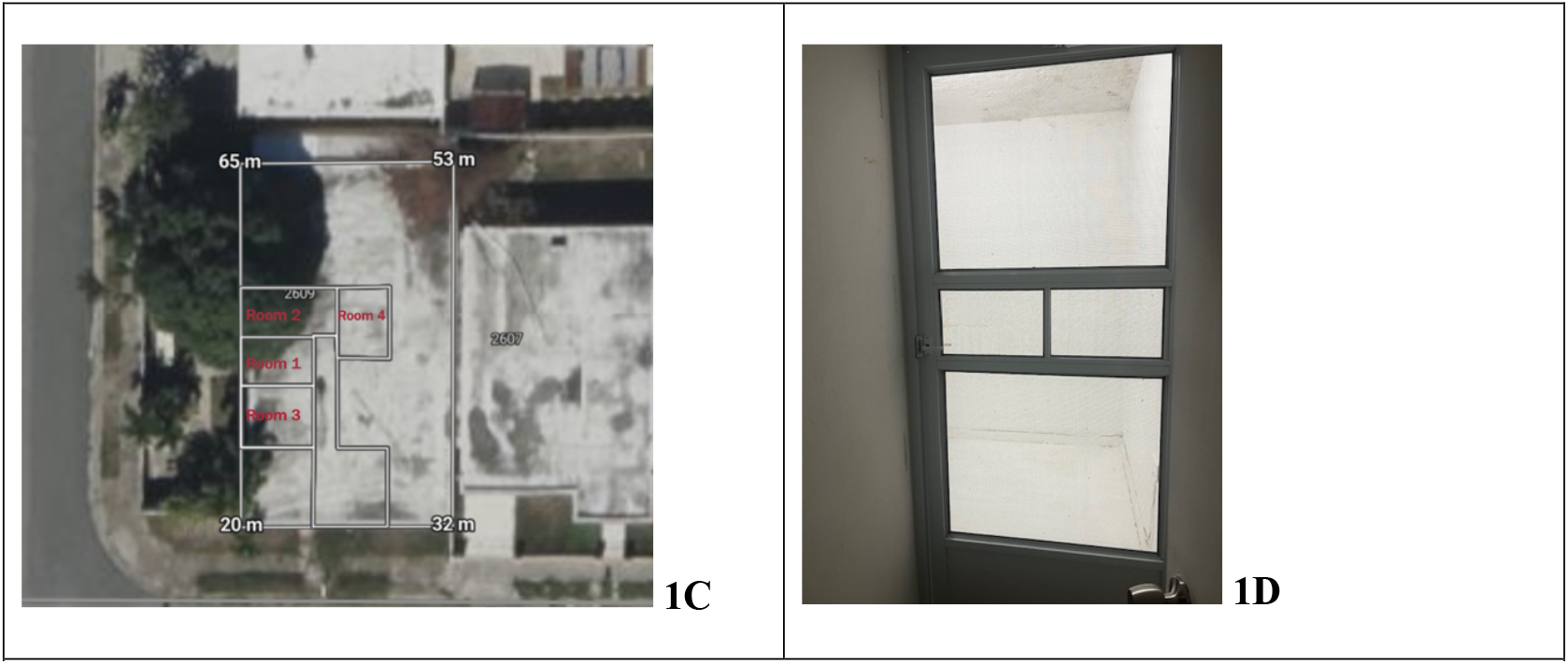
A, DABS foam drying overnight after soaking. Figure 1B, Cages used for laboratory cage trials. Figure 1C, Schematic layout of the experimental house used in semi field trials. Figure 1D, screen door placed outside each room, featuring a sliding panel for mosquito release; all interior walls are painted white.

#### DABS size experiment

To establish the optimal size of DABS, we compared DABS of different dimensions using both cages and experimental rooms. Four rooms and four rearing cages were treated with 1% DABS devices of varying sizes, small (¼ letter size, 150.9 cm^2^), medium (½ letter size, 301.9 cm^2^), and large (letter size, 603.8 cm^2^), along with a control medium sized DABS prepared using 10% sucrose solution without boric acid. 1% concentration was selected for trials due to previous data from Sippy et al. (2020). Mosquitoes were exposed to DABS for 48 hours, after which dead and alive specimens were counted. We performed four replicates of this experiment. Medium sized DABS are used for the rest of the trials.

#### DABS density experiment

To establish the optimal number of DABS per room, we compared rooms using one, two, and four 1% DABS, plus a control room containing four DABS manufactured using 10% sucrose solution without boric acid. Trials lasted 48 hours, and survival data was collected at the end. We performed four replicates of this experiment.

#### DABS light experiment

To establish the effect of light source in DABS efficiency, four rooms were divided between natural lighting conditions (open screened windows) and artificial lighting (ceiling lights with closed windows). Each room was tested using two medium sized 1% DABS and two control DABS without boric acid. Trials lasted 48 hours, and data was collected at the end. Four replicates of this design were performed throughout the trial period.

#### Release-recapture experiments

During the size, light, and density experiments, we observed increased ant activity in the experimental rooms, which posed a risk of interfering with mosquito recovery. As a result, all trials were paused for a period of nine months to allow for ant control measures. During this period, three rounds of insecticide treatments were conducted to address persistent ant infestations at the study site. Initial efforts involved over-the-counter products, including Sevin® Lawn Insect Killer (for the yard), Raid® Ant Baits, and Ortho® Home Defense (for indoor perimeters). These interventions were conducted in the month of November 2024, applied twice, two weeks apart. Despite these interventions, ant activity persisted the following month, prompting the engagement of a professional pest control service, Rentokil®. A commercial insecticide was applied inside and outside the house, but never in the experimental rooms to prevent confounding. Raid® Ant Baits were placed in the experimental rooms but were removed before any mosquito trials resumed. To further minimize the potential for residual insecticide effects on mosquito behavior or mortality, a defined waiting period was observed before initiating trials. December 2024, prior to restarting treatment trials, baseline recovery assessments were conducted to evaluate mosquito recapture efficiency in each of the four test rooms. For each baseline assessment, approximately 40 female Ae. aegypti mosquitoes were released per room, and after 24 hours, all mosquitoes were collected using a Prokopack Aspirator™ (John W. Hock Company®, Gainesville, Florida,USA). Recovered individuals were counted to determine recovery rates. These assessments were conducted once before the initial set of experimental house trials and repeated twice following the final pest treatment to confirm that ant presence had been effectively mitigated and would not confound the upcoming experiments.

#### DABS height trials

To establish the optimal height for DABS placement, each room received four 2% DABS devices placed at different heights: flushed to the ground, 0.75m, and 1.5m high, with a control room containing no DABS. These heights were selected based on previous studies showing that *Ae. aegypti* typically rest at heights below 1.5 meters, with one article reporting the highest concentrations occurring around the 0.2-meter mark inside homes (Dzul-Manzanilla et al., 2016; Facchinelli et al., 2023; Seang-arwut et al., 2023). This trial also used 2% concentration based on cage preliminary data indicating that this concentration might perform more rapidly than other doses in mortality. Trials lasted 48 hours, and data was collected at the end. These trials were replicated after ant treatment was done inside and around the house.

### Data analysis

For the cage trial datasets, time-to-event data were analyzed using survival analysis. Specifically, the Cox proportional hazards model was employed to assess differences in mosquito mortality over time across treatment groups. Survival objects were constructed using the Surv() function in the “survival” package in R (R Core Development Team, 2010; Therneau, 2024), incorporating both the time until death and censoring status. Proportional hazards assumptions were checked to ensure model validity.

Logistic regression was performed on the experimental house data to evaluate the effects of experimental conditions on mosquito survival outcomes as well as the recovery rates of mosquitoes. This analysis was conducted using generalized linear models (GLMs) with a binomial error distribution, implemented in R Studio version 4.5.1.

### Potential biases and treatment of the trial house

Modifying a vacant house into a semi-field environment introduced several uncontrollable factors that affected the trial results. Despite extensive attempts to achieve complete containment in the experimental rooms, it was not possible to fully prevent the incursion of insects and even some small reptiles from the surrounding environment. A particularly persistent issue was the presence of common house ants, which are widespread in Puerto Rico and frequently entered the space despite repeated mitigation efforts. These ants removed dead mosquitoes from the trial areas, carrying them back to their nests for food while trials were still underway (Figure 2). To address the ant problem, three rounds of insecticide were applied around the perimeter of the house and inside the living room. Two applications were performed using over the counter insecticides. Toward the end of the trials, a professional was hired to fumigate both the interior and exterior of the house. The individual trial rooms, however, were left untreated to avoid interference with the study outcomes.

**Figure 2:**
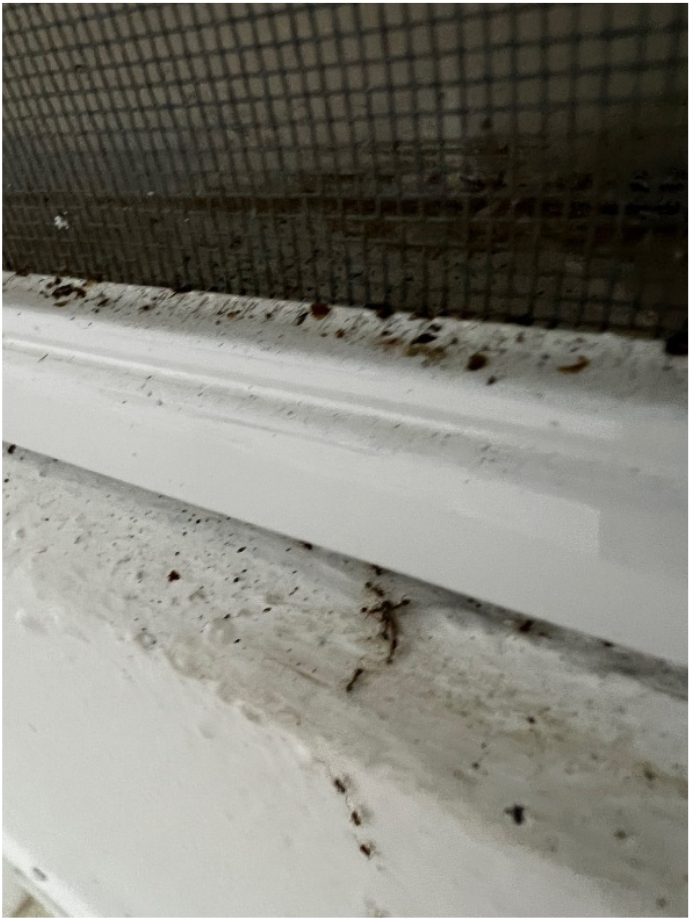
Ants carrying the remaining carcass of a dead mosquito toward their nest.

Screen doors were installed before the experimental period to ensure that mosquitoes were not escaping and that missing specimens had likely been removed post-mortem by ants rather than exiting the rooms. Environmental modifications, such as the installation of screen doors, may have also influenced mosquito behavior or altered handling protocols, potentially affecting methodological consistency and result reliability. Additionally, the study was conducted by multiple individuals over time, which may have introduced variability in trial execution and data collection.

## Results

### Cage trials

After 48-hours exposure to DABS, *Ae. aegypti* in cage trials suffered nearly complete mortality compared to control cages which had near universal survival. All tested concentrations resulted in significantly higher mortality compared to controls (Cox proportional hazards model: p < 0.001), Figure 3)(Figure 6). However, no significant differences in mortality were observed among the DABS-treated groups (Wald test: p = 0.50) (Figure 3A). The availability of water in the cages attenuated the effect of DABS, reducing the mortality by 47% in females (Figure 3B, Cox proportional hazards model: p < 0.001) and 32% in males (Figure 3C, Cox proportional hazards model: p < 0.001).

**Figure 3.**
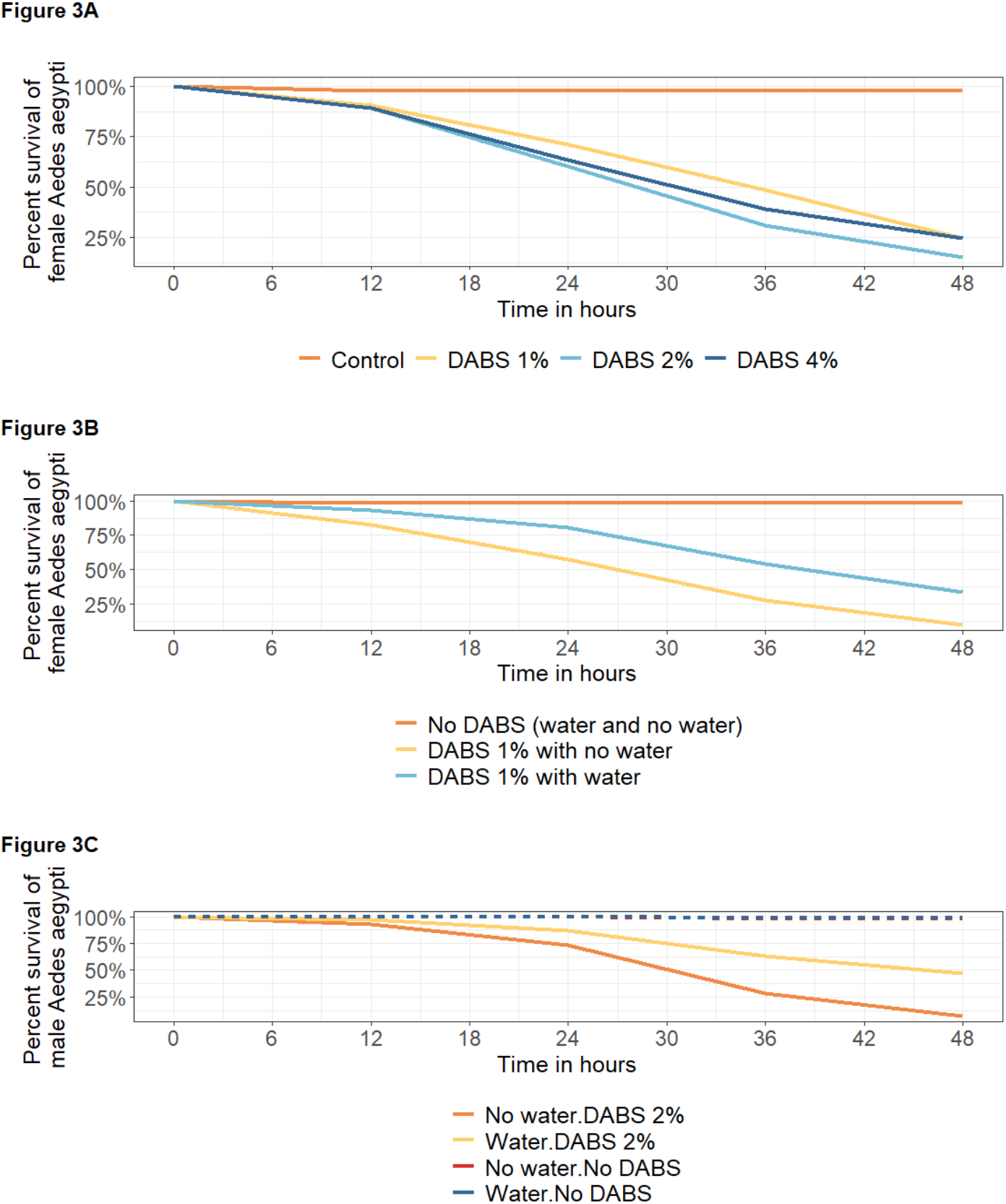
The survival outcomes of *Ae. aegypti* in cage trials, data collected every 6 hours. Panel A varies the concentration of boric acid applied of the DABS (1%, 2%, and 4%) and mortality in female *Ae. aegypti*. Panels B and C show the effect of DABS in the presence of water or not, for female and male *Ae. aegypti*. Panel C, both variables of “No water.No DABS” and “Water.No DABS” are overlapping in the figure.

### Experimental house

Across all semi-field trials, no significant differences in recovery were observed between rooms when comparing the control arm of the experiments (binomial GLM, Room 2, p = 0.659; Room 3, p = 0.860; Room 4, p = 0.158; reference = Room 1) (Figure 4A). However, when experimental condition (density, size, height, light) was included in the regression model, recovery in Room 2 was significantly lower (binomial GLM, p = 0.0124), an effect primarily driven by the electric light experiment (binomial GLM, p = 0.0149). There is also a slightly better recovery of *Aedes aegypti* following pest control interventions, although the difference is not substantial (Figure 4B & 4C).

**Figure 4:**
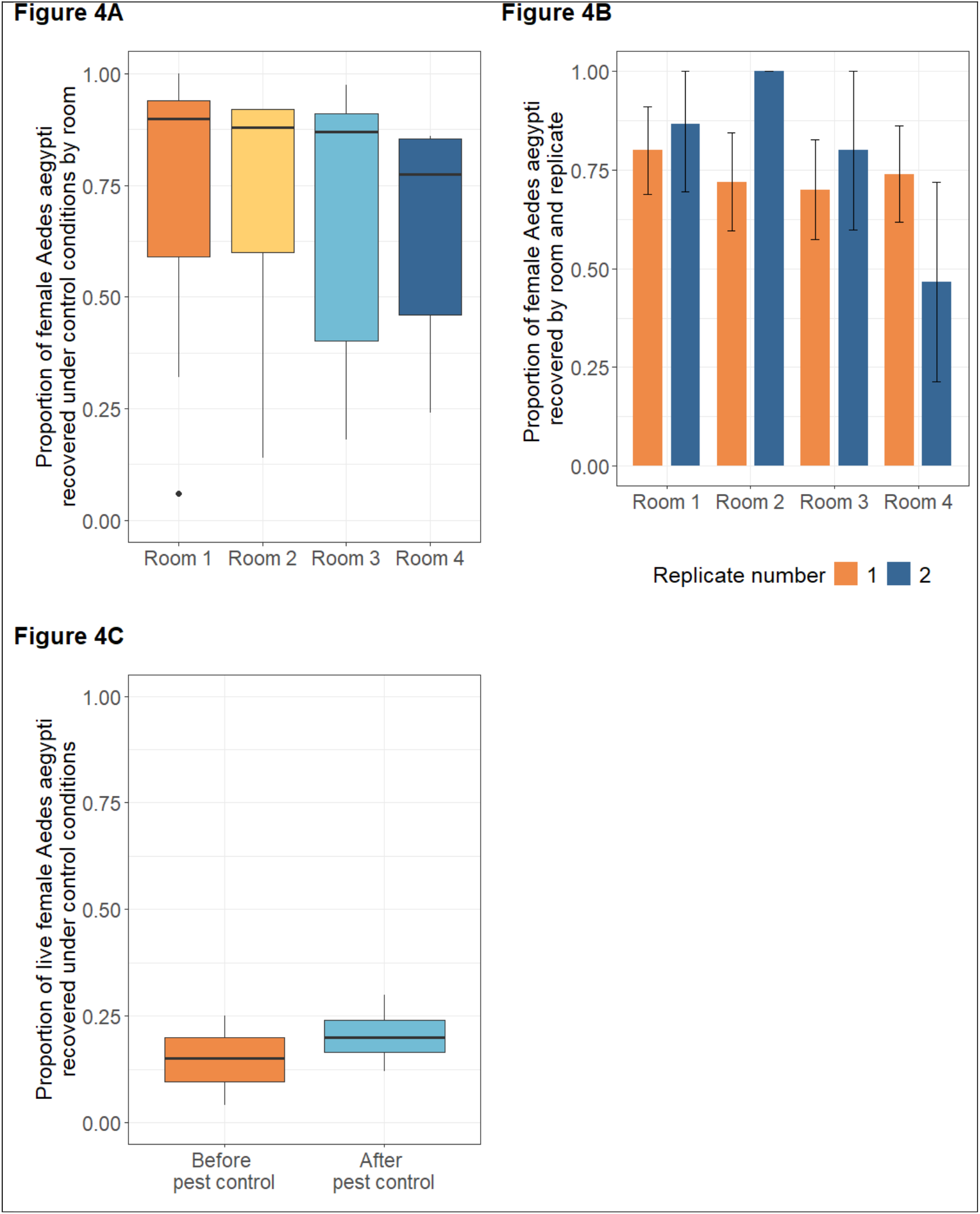
Recovery of *Ae. aegypti* in semi field trials after 48 hours. Panel A shows the proportion of *Ae. aegypti* females recovered per room under control conditions during house trials. Panel B illustrates female *Ae. aegypti* recovery by room before and after pest control interventions. Panel C illustrates female *Ae. aegypti* recovery under control conditions, before and after pest control treatment.

In experimental house trials (Figure 5), DABS treatments significantly reduced survival compared to controls, demonstrating ∼75% effectiveness (binomial GLM: one DABS, *p* < 0.001; two DABS, *p* < 0.001; four DABS, *p* < 0.001). Deploying one or two devices resulted in marginally higher mortality than four devices, the differences were suggestive but not statistically significant (*p* = 0.06 and *p* = 0.10, respectively) (Figure 5A).

**Figure 5:**
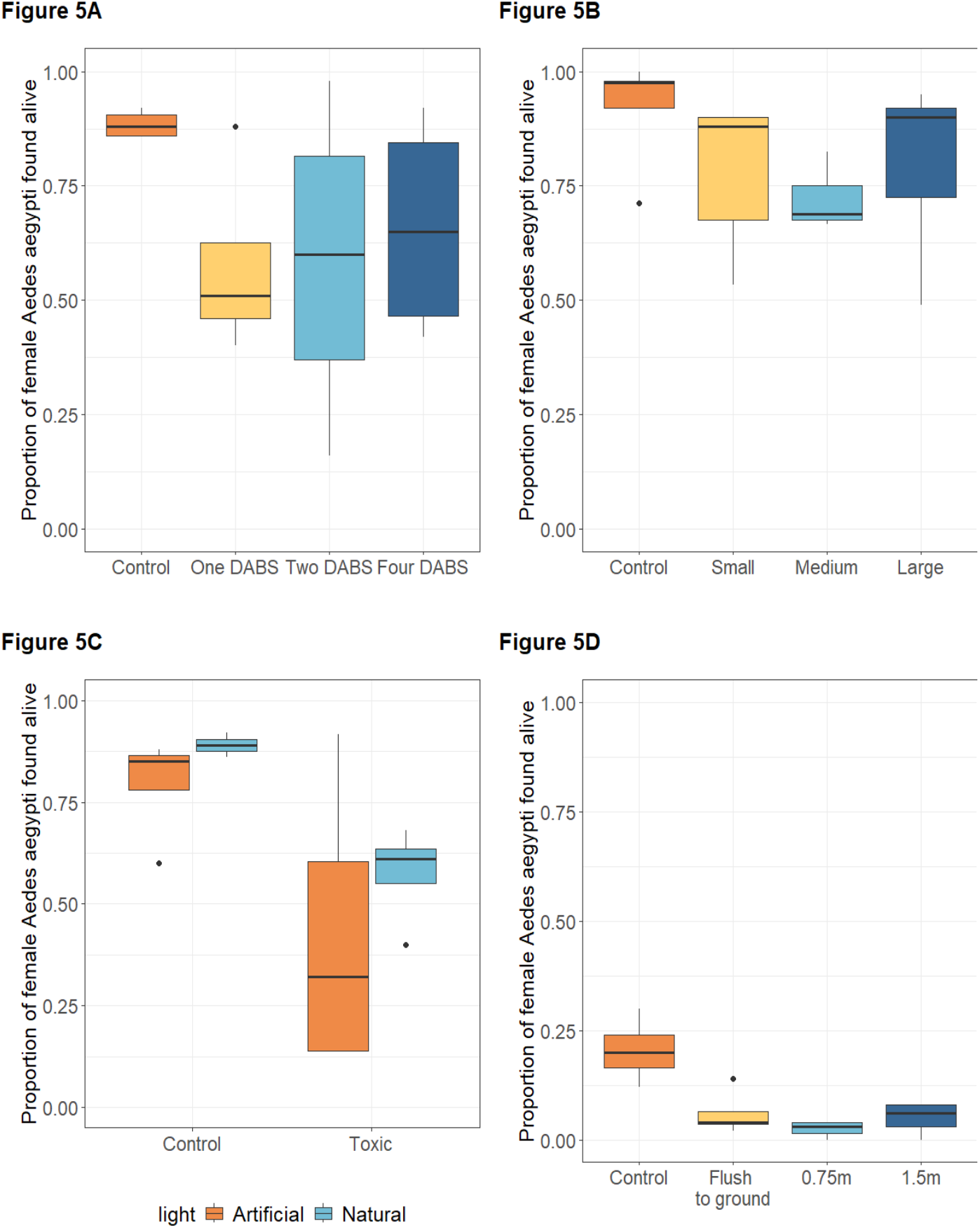
Illustrates the recovery after 48 hours of female *Ae. aegypti* in house trials using DABS devices with varying density, light, size, and height to analyze their effectiveness. Panel A illustrates the proportion of live *Ae. aegypti* females recovered in house trials across treatments with varying numbers of DABS devices. Panel B shows the impact of DABS device size on *Ae. aegypti* survival. Panel C presents the proportion of live mosquitoes recovered under different lighting conditions (natural vs. artificial light) during DABS exposure. Panel D compares *Ae. aegypti* survival across different DABS installation heights.

All tested DABS sizes significantly reduced survival relative to controls (small, *p* < 0.001; medium, *p* < 0.001; large, *p* = 0.0023) (Figure 5B & 6). No significant differences in mosquito survival were found between the three tested DABS sizes, with marginal better performance of medium-sized devices over large ones (p=0.09). While natural light alone was associated with higher mosquito survival (*p* = 0.028), the interaction between light and DABS treatment was not significant (*p* = 0.322) (Figure 5C). All tested device heights significantly reduced survival compared to controls (*p* < 0.001), with devices placed at 0.75 m being marginally more effective than those flush to the ground (*p* = 0.08), and similar in performance to those at 1.5 m (*p* = 0.18) (Figure 5D).

### Supplementary Table

**Figure 6**

#### Summary of Cox PH models results

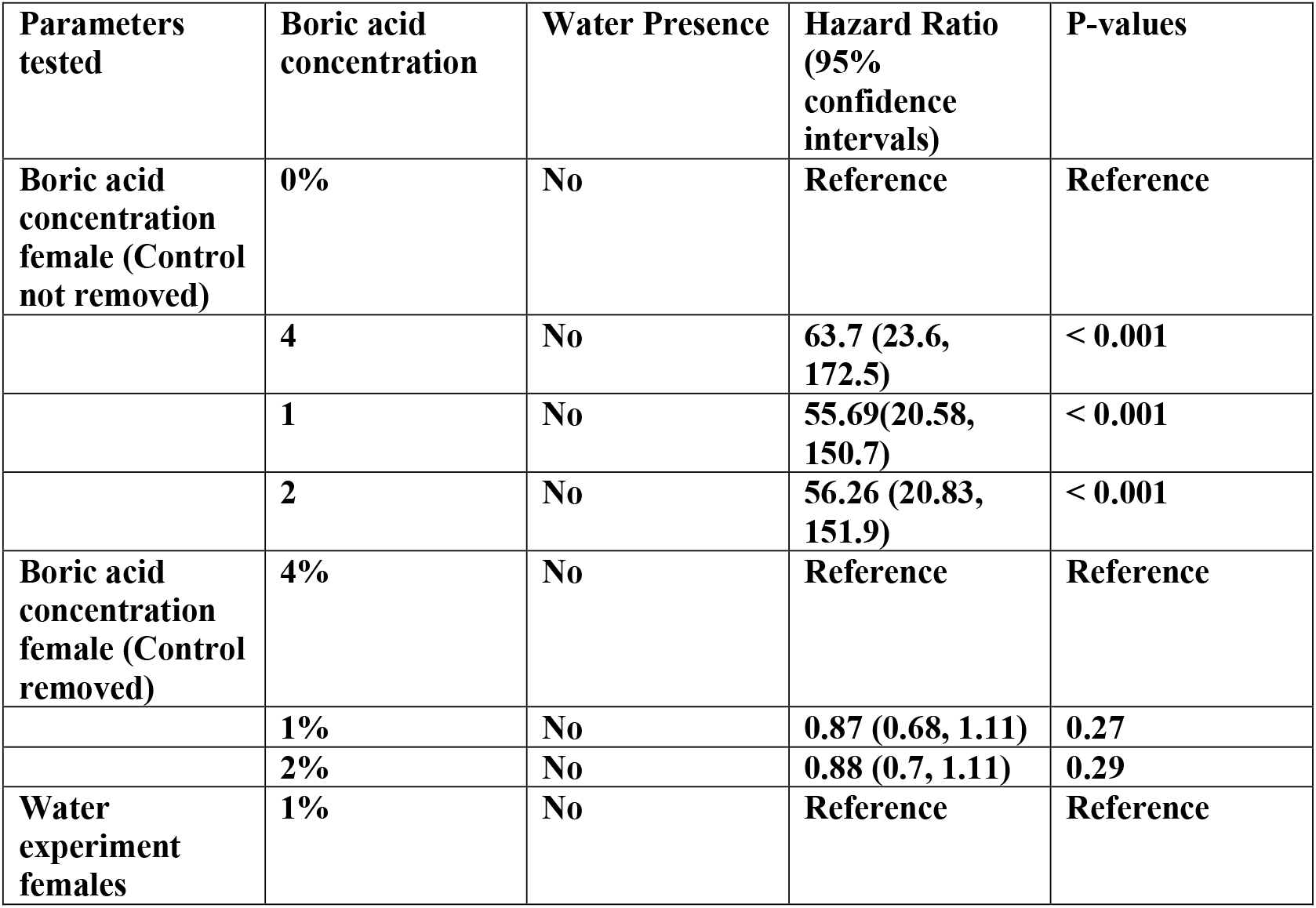
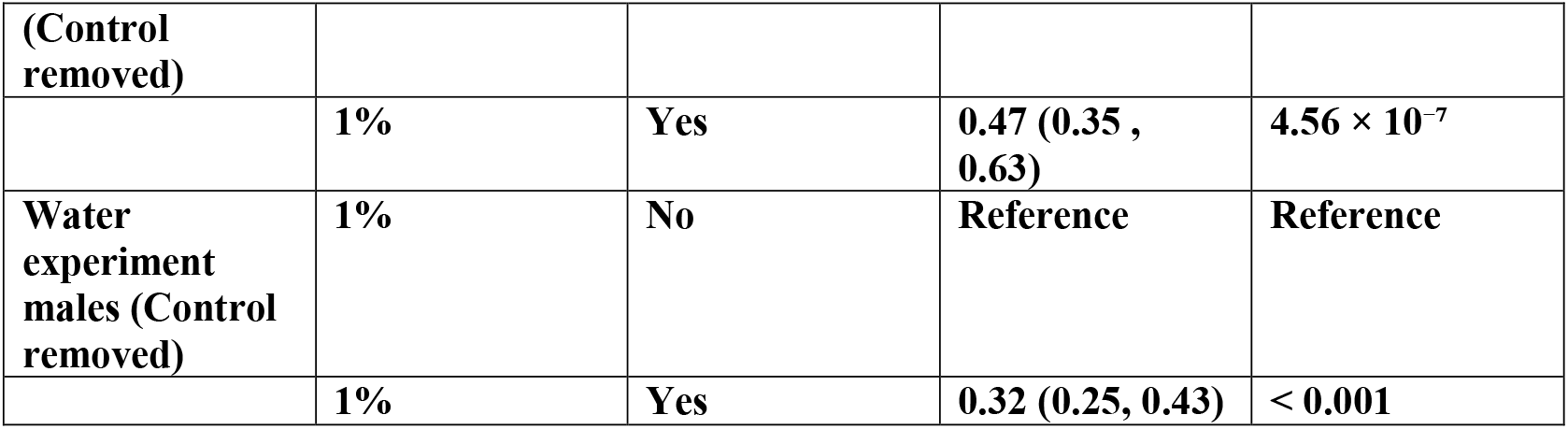

## Discussion

The cage trials demonstrated a strong lethal effect of DABS *Ae. aegypti* mortality. Notably, investigations into the concentration of boric acid in these laboratory settings revealed that increasing the concentration beyond 1% did not result in additional lethality. Lower concentrations may be preferable to reduce cost and environmental impact without sacrificing effectiveness. The presence of water was found to substantially interfere with the ingestion-based toxicity mechanism in both male and female *Ae. aegypti*. As a result, the efficacy of DABs in real-world settings may be reduced by the availability of water sources. Further field evaluations are necessary to assess the magnitude of this effect in the field.

Experimental house trials provide additional valuable data that can inform future deployment strategies. Density trials resulted in marginally higher mortality with one or two DABS devices compared to four, yet due to the lack of statistical significance, there is no definitive advantage to using fewer DABS per room in our setting. Similarly, trials evaluating device size resulted in no significant difference in mosquito survival between the three different sizes of DABS, with marginal better performance of medium DABS when compared to large DABS. Both factors need to be further studied for the manufacturing of the DABS device. Natural lighting without supplemental electric lighting appeared to attenuate, but not eliminate, the killing effect of DABS. This finding is important when considering the diversity of indoor environments in different communities. Other studies have found that *Ae. aegypti* tend to inhabit dark, secluded indoor spaces such as restrooms and closets where humidity is high and light disturbance is minimal (Seang-arwut et al., 2023). The trials to identify the optimal height for DABS placement provided useful operational guidance: DABS placed flush to the ground were less effective, indicating that higher placement around 0.75m-1.5m, beyond the reach of crawling children and small pets, is both safer and more effective.

The observed results in these trials are consistent with previous preliminary studies on boric acid-treated ATSBs, which examined various concentrations of boric acid and how age and sex affected *Ae. aegypti* mortality. Those studies identified 1% boric acid as the optimal concentration, validating our finding of no significant increase in mortality beyond this concentration, and found newly emerged adults to be more susceptible than older specimens (Hossain et al., 2021).

Interestingly, this study also reported significant differences in the lethal concentration (LC_50_) between male and female *Ae. aegypti*, ranging from 0.54 to 20.92, with LC_50_ values decreasing between 48 and 72 hours. In contrast, our study observed near-complete lethality within 48 hours of exposure to DABS in both male and female cage trials, with no significant difference between sexes. Similarly, Sippy et al., 2020 reported complete lethality under controlled conditions within 48 hours and higher mosquito mortality in treated rooms compared to controls (Sippy et al., 2020), findings consistent with ours in both cage and semi-field trials. While some of these studies used different attractants or liquid sugar baits, our results confirm that *Ae. aegypti* can be reliably killed using boric acid in a dry bait format. These results underscore the potential of dry bait systems and contribute to growing evidence supporting DABS.

While these results are promising, several challenges encountered during the household trials introduced potential sources of bias. A significant limitation emerged in the form of ant interference, which led to the removal of mosquito carcasses and introduced potential data loss. While targeted insecticide applications slightly reduced the ant population, the interference of ants in the experimental house raises important questions and uncertainties. We are unsure whether the ants were attracted to the sugar from the DABS or the high amount of mosquito carcasses during each trial. Future formulations may benefit from understanding the attraction from different sugar sources. For example, other ATSBs have been successful with a variety of sugar and fresh fruit juices, results showing higher efficacy with guava, plum, and mango juice (Kumar et al., 2022). Yet it is unknown if fruit-based sugar sources might be more attractive to ants compared to regular cane sugar used in our trials. Despite these complications, consistent baseline recovery across the different rooms under control conditions suggests that the study design was robust and yielded repeatable results.

Based on current findings, the next steps for advancing this novel intervention include field validation, optimization of the sweetener attractant, assessment of non-target effects on indoor pests, and evaluation of community acceptance and feasibility of implementation. Given the variability of environmental conditions and the need for additional validation, expanded field trials are warranted. These trials will support the refinement of DABS design and deployment protocols, ultimately enhancing their reliability, efficacy, and scalability for mosquito control across diverse ecological settings.

## Data Availability

All data produced in the present study are available upon reasonable request to the authors

## Acknowledgements

We thank Marta I. Diaz-Garcia, Luis Rivera, Herick Leon, Hernan Olivera, Damian Suren, Raiza Alvarado, and Joy Adkins for their contributions to the project. This work was supported by a broad agency agreement with the CDC, contract number BAA 75D30122C15613. The findings and conclusions in this report are those of the authors and do not necessarily represent the views of the CDC.

## References

CDC. (2025). Current Dengue Outbreak. Centers for Disease Control and Prevention. https://www.cdc.gov/dengue/outbreaks/2024/index.html

CFWSC. (2016). Climate of Puerto Rico. Caribbean-Florida Water Science Center (CFWSC). https://www.usgs.gov/centers/cfwsc/science/climate-puerto-rico?utm_source=chatgpt.com

CIMH. (2014). Ponce Rainfall. Caribbean Regional Climate Center.

Cochran, D. G. (1995). Toxic effects of boric acid on the German cockroach. Experientia, 51(6), 561–563. 10.1007/BF02128743

DATAUSA. (2023). Ponce, PR Metropolitans Statistical Area. Data USA. https://datausa.io/profile/geo/ponce-pr-31000US38660

Dennis, E. J., Goldman, O. V., & Vosshall, L. B. (2019). Aedes aegypti Mosquitoes Use Their Legs to Sense DEET on Contact. Current Biology, 29(9), Article 9. 10.1016/j.cub.2019.04.004

Dusfour, I., Vontas, J., David, J.-P., Weetman, D., Fonseca, D. M., Corbel, V., Raghavendra, K., Coulibaly, M. B., Martins, A. J., Kasai, S., & Chandre, F. (2019). Management of insecticide resistance in the major Aedes vectors of arboviruses: Advances and challenges. PLoS Neglected Tropical Diseases, 13(10), e0007615. 10.1371/journal.pntd.0007615

Dzul-Manzanilla, F., Ibarra-López, J., Bibiano Marín, W., Martini-Jaimes, A., Leyva, J. T., Correa-Morales, F., Huerta, H., Manrique-Saide, P., & Prokopec, G. V. M. (2016). Indoor Resting Behavior ofAedes aegypti(Diptera: Culicidae) in Acapulco, Mexico. Journal of Medical Entomology, tjw203. 10.1093/jme/tjw203

EPA. (2025). Biopesticides Active Ingredients. Ingredients Used in Pesticide Products. https://www.epa.gov/ingredients-used-pesticide-products/biopesticide-active-ingredients?utm_source=chatgpt.com

Estep, A. S., Sanscrainte, N. D., Waits, C. M., Louton, J. E., & Becnel, J. J. (2017). Resistance Status and Resistance Mechanisms in a Strain of Aedes aegypti (Diptera: Culicidae) From Puerto Rico. Journal of Medical Entomology, 54(6), 1643–1648. 10.1093/jme/tjx143

Facchinelli, L., Alsharif, B., Jones, J. D., Matope, A., Barbosa, R. M. R., Ayres, C. F. J., & McCall, P. J. (2023). Mapping Aedes aegypti indoor resting behavior reveals a preference vulnerable to householder-led vector control. PNAS Nexus, 2(7). 10.1093/pnasnexus/pgad226

Fiorenzano, J. M., Koehler, P. G., & Xue, R.-D. (2017). Attractive Toxic Sugar Bait (ATSB) For Control of Mosquitoes and Its Impact on Non-Target Organisms: A Review. International Journal of Environmental Research and Public Health, 14(4), 398. 10.3390/ijerph14040398

Grabenstein, J. D., & Tomar, A. S. (2023). Global geotemporal distribution of chikungunya disease, 2011–2022. Travel Medicine and Infectious Disease, 54, 102603. 10.1016/j.tmaid.2023.102603

Habes, D., Morakchi, S., Aribi, N., Farine, J.-P., & Soltani, N. (2006). Boric acid toxicity to the German cockroach, Blattella germanica: Alterations in midgut structure, and acetylcholinesterase and glutathione S-transferase activity. Pesticide Biochemistry and Physiology, 84(1), 17–24. 10.1016/j.pestbp.2005.05.002

Hossain, Md. F., Ghosh, A., Hossain, M. A., & Seheli, K. (2021). Efficacy of boric acid as an attractive toxic sugar bait on laboratory reared Aedes aegypti Linnaeus (Diptera: Culicidae). International Journal of Mosquito Research, 8(4), 47–52. 10.22271/23487941.2021.v8.i4a.553

Khallaayoune, K., Qualls, W. A., Revay, E. E., Allan, S. A., Arheart, K. L., Kravchenko, V. D., Xue, R.-D., Schlein, Y., Beier, J. C., & Müller, G. C. (2013). Attractive Toxic Sugar Baits: Control of Mosquitoes With the Low-Risk Active Ingredient Dinotefuran and Potential Impacts on Nontarget Organisms in Morocco. Environmental Entomology, 42(5), 1040–1045. 10.1603/EN13119

Kumar, S., Sharma, A., Samal, R. R., Kumar, M., Verma, V., Sagar, R. K., Singh, S., & Raghavendra, K. (2022). Attractive Sugar Bait Formulation for Development of Attractive Toxic Sugar Bait for Control of Aedes aegypti (Linnaeus). Journal of Tropical Medicine, 2022, 1–10. 10.1155/2022/2977454

Murray, F. J. (1995). A Human Health Risk Assessment of Boron (Boric Acid and Borax) in Drinking Water. Regulatory Toxicology and Pharmacology, 22(3), 221–230. 10.1006/rtph.1995.0004

Noisumdaeng, P., Dangsagul, W., Sangsiriwut, K., Prasertsopon, J., Changsom, D., Yoksan, S., Ajawatanawong, P., Buathong, R., & Puthavathana, P. (2023). Molecular characterization and geographical distribution of Zika virus worldwide from 1947 to 2022. International Journal of Infectious Diseases, 136, 5–10. 10.1016/j.ijid.2023.08.023

Paeporn, P., Supaphathom, K., Sathantriphop, S., Mukkhun, P., Sangkitporn, S., & New, D. (2005). Insecticide Susceptibility of Aedes aegypti in Tsunami-affected Areas in Thailand. (Dengue Bulletin.). World Health Organization. https://iris.who.int/bitstream/handle/10665/164109/dbv29p210.pdf?sequence=1

Pearson, M. A., Blore, K., Efstathion, C., Aryaprema, V. S., Muller, G. C., Xue, R., & Qualls, W. A. (2020). Evaluation of boric acid as toxic sugar bait against resistant Aedes aegypti mosquitoes. Journal of Vector Ecology, 45(1), 100–103. 10.1111/jvec.12377

Powell, J. R., & Tabachnick, W. J. (2013). History of domestication and spread of Aedes aegypti—A Review. Memórias Do Instituto Oswaldo Cruz, 108(Suppl 1), 11–17. 10.1590/0074-0276130395

R Core Development Team. (2010). R: A Language and Environment for Statistical Computing. Http://Www.R-Project.Org/.

Rodríguez, M. M., Bisset, J. A., & Fernández, D. (2007). LEVELS OF INSECTICIDE RESISTANCE AND RESISTANCE MECHANISMS IN AEDES AEGYPTI FROM SOME LATIN AMERICAN COUNTRIES. Journal of the American Mosquito Control Association, 23(4), 420–429. 10.2987/5588.1

Seang-arwut, C., Hanboonsong, Y., Muenworn, V., Rocklöv, J., Haque, U., Ekalaksananan, T., Paul, R. E., & Overgaard, H. J. (2023). Indoor resting behavior of Aedes aegypti (Diptera: Culicidae) in northeastern Thailand. Parasites & Vectors, 16(1). 10.1186/s13071-023-05746-9

Sippy, R., Rivera, G. E., Sanchez, V., Heras, F., Morejón, B., Beltrán, E., Hikida, R. S., López-Latorre, M. A., Aguirre, A., Stewart-Ibarra, A. M., Larsen, D. A., & Neira, M. (2020). Ingested insecticide to control Aedes aegypti: Developing a novel dried attractive toxic sugar bait device for intra-domiciliary control. Parasites & Vectors, 13(1), 78. 10.1186/s13071-020-3930-9

Therneau, T. M. (2024). A Package for Survival Analysis in R. https://CRAN.R-project.org/package=survival

United States Census Bureau. (2020). Ponce Municipio, Puerto Rico. United States Census Bureau. https://data.census.gov/profile/Ponce_Municipio,_Puerto_Rico?g=050XX00US72113#po pulations-and-people

USAID. (2019). OPERATION OF AN INSECTARY PRACTICAL MANUAL AEDES AEGYPTI REARING PROCEDURES AND BASIC PRINCIPLES OF BIOSAFETY. United States Agency for International Development. https://api.globalvectorhub.lshtm.ac.uk/resource/serve-file/184/5ebae58d4619b_12.%20ZAP%20insectary%20management+Ae%20aegypti%20rearing.pdf

Ware-Gilmore, F., Rodriguez, D. M., Ryff Mph, K., Torres, J. M., Velez, M. P., Torres-Toro, C. T., Santiago, G. A., Rivera, A., Madewell, Z. J., Maldonado, Y., Cardona-Gerena, I., Brown, G. C., Adams, L. E., Paz-Bailey, G., & Marzán-Rodriguez, M. (2025). Dengue Outbreak and Response—Puerto Rico, 2024. MMWR. Morbidity and Mortality Weekly Report, 74(5), 54–60. 10.15585/mmwr.mm7405a1

WHO. (n.d.). Improving data for dengue. World Health Organization. https://www.who.int/activities/improving-data-for-dengue

WHO. (2024). Dengue and severe dengue. World Health Organization. https://www.who.int/news-room/fact-sheets/detail/dengue-and-severe-dengue

